# Leveraging the electronic health record to identify delivery of goal-concordant care

**DOI:** 10.1101/2024.09.24.24314226

**Authors:** Catherine L Auriemma, Anne Song, Lake Walsh, Jason Han, Sophia Yapalater, Alexander Bain, Lindsay Haines, Stefania Scott, Casey Whitman, Stephanie P Taylor, Gary E. Weissman, Matthew J Gonzales, Roshanthi Weerasinghe, Staci J Wendt, Katherine R Courtright

## Abstract

**Background:** Goal-concordant care (GCC) is recognized as the highest quality of care and most important outcome measure for serious illness research, yet there is no agreed-upon or validated method to measure it.

**Objective:** Assess feasibility of measuring GCC using clinical documentation in the electronic health record (EHR).

**Design:** Retrospective chart review study.

**Participants:** Adults with ≥50% predicted six-month mortality risk admitted to three urban hospitals in a single health system. All participants had goals-of-care (GOC) discussions documented in the EHR 6 months before and 6 months after admission manually classified into one of four categories of goals: (1) comfort-focused, (2) maintain or improve function, (3) life-extension, or (4) unclear.

**Main Measures:** Pairs of physician-coders independently reviewed EHR notes from 6 months before through 6 months after admission to identify and classify care received between each documented GOC discussion into one of the four goals categories. Epochs between GOC discussions were then coded as goal-concordant if GOC and care received classifications were aligned, goal-discordant if they were misaligned, or uncertain if either classification was unclear or not documented. Coder inter-rater reliability was assessed using kappa statistics.

**Key Results:** Inter-rater reliability for classifying care received was almost perfect (95% interrater agreement; Cohen’s kappa=0.92; 95% CI, 0.86-0.99). Of 398 total epochs across 109 unique patients, 198 (50%) were goal-concordant, 112 (28%) were of uncertain concordance, and 88 (22%) were goal-discordant. Eighty (73%) patients received care of uncertain concordance during at least one epoch. Forty-eight (44%) patients received goal-discordant care during at least one epoch.

**Conclusions:** Clinician chart review was a feasible method for measuring GCC and can inform natural language processing and machine learning methods to improve the clinical and research utility of this method. More work is needed to understand the driving factors underlying the high rate of uncertain concordance and goal-discordant care identified among this seriously ill cohort.

## 1 Introduction

While goal-concordant care (GCC) is widely considered the highest quality of care,^1,2^ there is no agreed-upon or validated method to measure it.^3–6^ GCC is defined as medical care that aligns with and promotes patients’ goals and preferences regarding treatment intensity, functional outcomes, and longevity. Delivery of GCC is recognized as a key priority of numerous medical specialties and societies^7–10^ and has been identified as the most important outcome measure for assessments of serious illness care, advance care planning (ACP), and studies of palliative and end-of-life care research.^11^ However, the inability to reliably measure it impedes efforts to develop and test interventions aimed at increasing GCC.

Prior approaches to measuring GCC have focused on proxy assessments of communication^12^ or narrowly defined populations, such as advanced cancer.^3,13,14^ Patient-, caregiver-, or clinician-reported assessments^15,16^ suffer from desirability, recall, and selection biases, and are prone to data missingness. Furthermore, collection of these assessments is often infeasible at scale. A framework for measuring GCC using clinical notes in the electronic health record (EHR)^5^ has the advantages of allowing for longitudinal assessment, using a data source that contains a wealth of information on care delivery, and is already the location that is considered best practice for real-world goals-of-care (GOC) conversations to be documented.^1^

Despite efforts to promote systematic documentation of patients’ GOC in clearly delineated locations within the EHR, adoption of standardized tools and note templates is suboptimal. Indeed, a substantial proportion of GOC conversations are documented outside of designated places within the EHR.^17–20^ However, as artificial intelligence methods continue to advance and enable increasingly accurate approaches to analyze free-text clinical data,^17,18,21–25^ a feasible and pragmatic approach to measure GCC using EHR data may be within reach. In this retrospective cohort study, we sought to operationalize an adapted framework^5^ to identify and measure GCC through manual chart review.

## 2 Methods

### 2.1 Study Design and Setting

We conducted a retrospective chart review study among 109 seriously ill patients admitted to one of three urban, academic-affiliated hospitals in Philadelphia, Pennsylvania within the University of Pennsylvania Health System (UPHS) between April 1, 2019 and July 31, 2019. The Institutional Review Board at the University of Pennsylvania approved this study with a waiver of informed consent.

### 2.2 Study Population

Details of this study population were published previously.^20^ Briefly, we identified all adults (≥18 years of age) hospitalized for ≥3 calendar days during the study period, with ≥50% predicted risk of six-month mortality at admission,^26^ and at least one prior inpatient or outpatient encounter within the study health system in the 12 months preceding the enrollment hospital admission. The sample was stratified by six-month mortality risk (high 50-74% and very high ≥75%) and then randomly sampled to obtain the final cohort. Finally, as described in our prior work and detailed in Figure 1, we identified all GOC discussions^27^ that were documented during inpatient, ambulatory, and home-care encounters six months before and after the index admission and classified the patients’ goals into four categories: (1) comfort-focused, (2) maintain or improve function, (3) life-extension, or (4) unclear. If no GOC discussion was documented in the six months preceding the index admission, baseline goals were categorized as unclear.

**Figure 1.**
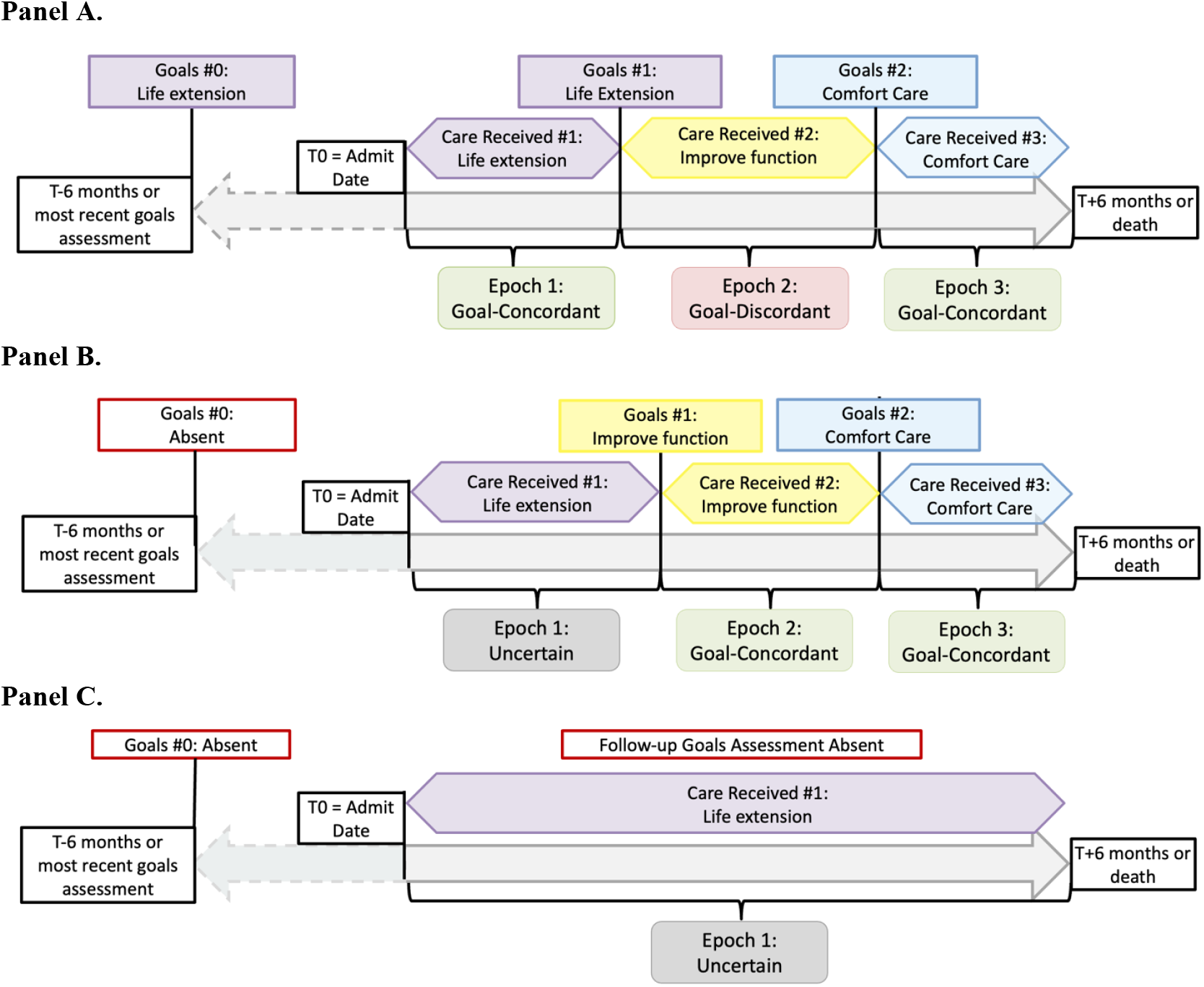
Assessments of goals-of-care (GOC) and care received over time illustrating delineation of epochs and identification of goal-concordant, goal-discordant, and uncertain care. **Panel A**. Patient record with documented GOC discussions in the 6 months prior to enrollment date and during follow-up. **Panel B**. Patient record with no documented GOC in the 6 months prior to enrollment date but multiple GOC assessments during follow-up. **Panel C**. Patient record with no documented GOC in the 6 months prior to enrollment date or during follow-up.

### 2.3 Data Collection and Variable Definitions

#### 2.3.1 Patient Characteristics and Clinical Data

Patients’ sociodemographic data were obtained from the UPHS EHR clinical data warehouse: self-reported binary sex, race, ethnicity, religion, and primary language. An inpatient palliative care consult was identified by the presence of a signed order during the index hospital encounter. We additionally collected readmissions to any UPHS hospital, home care visits, and subsequent referrals to home or outpatient palliative care during the six-month follow-up period.

#### 2.3.2 Care Received Classification

We assessed care received longitudinally from the date of index hospital admission through death or six months of follow-up, whichever was shorter. The six-month follow-up period was further divided into epochs between each identified GOC discussion (Figure 1). Each epoch was reviewed by a pair of reviewers blinded to the GOC categorizations to determine the category of care received during that time. All advance care planning notes, palliative care consult notes, and other clinical notes in which a GOC discussion had been identified were excluded from the care received assessment so as not to influence the reviewers’ classifications.

Reviewers were instructed to classify the perceived intention of the medical care received during each epoch through clinical text review. Care received was categorized using the same four categories as GOC (Table 1). “Life extension” included therapies delivered with the primary goal to prolong life, regardless of whether treatments might exacerbate pain, discomfort, or dysfunction in the short- or long-term. “Maintain or improve function” included therapies delivered with the goal of promoting the patient’s physical or cognitive function, even if treatments might exacerbate pain, discomfort, or dysfunction in the short-term. “Comfort-focused” care included therapies with the goal of relieving or avoiding suffering, including withholding or withdrawing life-prolonging therapies.

**Table 1.** Care received classification, definitions, and example descriptions from data abstraction, including excerpts from the electronic health record.

| Category | Definition | Example Descriptions from Data Abstractors |
| --- | --- | --- |
| Life extension | Care plan focuses on maximizing the patient’s longevity without limitations on care. Extending longevity or survival are prioritized over maximizing function or comfort. | Direct admission for kidney transplant (under general anesthesia, intubation, central line, arterial line), also underwent peritoneal dialysis catheter removal intra-operatively. Extubated post-operatively prior to transfer to floor, eventually discharged home. |
|  |  | Metastatic breast cancer complicated by malignant effusions, ventilator-dependent respiratory failure secondary to pneumonia. Underwent tracheostomy and feeding tube placement. Continued on vasopressors, total enteral nutrition, and ventilator wean. Readmitted for multifactorial respiratory failure to acute care hospital, then transferred to long-term acute care hospital. |
|  |  | Decompensated cirrhosis admitted with AKI. Previously on transplant list, deactivated on admission temporarily given functional decline. Nasogastric tube placed for supplemental enteral nutrition. Developed worsening labs concerning for infection, started antibiotics. Underwent diagnostic/therapeutic paracentesis and thoracentesis. Admitted to medical ICU for worsening respiratory failure on noninvasive ventilation, then intubated. Goal of stabilization in order to consider living donor liver transplant. |
| Maintain or improve function | Care plan focuses on maintaining or improving cognitive or physical function by preventing or reversing dysfunction, even if that medical care would increase discomfort. However, care that would increase survival/longevity without preservation or improvement in function is generally avoided. | Admitted to neuro ICU for acute ischemic stroke, underwent carotid endarterectomy; transferred post-op to ICU for frequent neuro checks; arterial line continued. Continued chronic intermittent hemodialysis. Discharged to acute rehab; readmitted due to anemia concerning for gastrointestinal bleed complicated by non-ST elevation myocardial infarction. Patient stated DNR/DNI. Underwent esophagogastroduodenoscopy (under monitored anesthesia care), which found 2 arterio-venous malformations, treated with argon plasma coagulation; discharged again to acute rehab. |
|  |  | Admitted to acute rehab. Patient reported to provider she would like time-limited trial of intubation, so code status changed to full code. Improved activities of daily living with physical, occupational, and speech therapy and discharged home. Underwent outpatient colonoscopy (under monitored anesthesia care). Underwent outpatient interventional radiology arterio-venous fistulogram; Rapid response called for hypertension, chest pain, nausea, and transferred to ED, then discharged home. |
|  |  | Patient continued on ventilator via tracheostomy. Goal of treating pain and anxiety without invasive procedures. Code status switched [from full code] to DNR, may intubate. Not yet ready for palliative ventilator liberation, wants to go to vent-capable facility and maximize quality of life. Readmitted to medical ICU with respiratory failure, started on antibiotics. Air hunger was treated with morphine and remained on mechanical ventilation throughout admission. Discharged to skilled nursing facility. |
| Comfort-focused | Care plan focuses on maximizing comfort and relieving or avoiding suffering. Includes seeking interventions to promote comfort and avoiding interventions that would increase discomfort, even at the expense of decreasing longevity. | PICC line placed. Code status changed to DNR/DNI. Tachyarrhythmia treatments disabled from pacemaker and implanted defibrillator. Continued on palliative milrinone. Discharged to home hospice. |
|  |  | Code status changed to “DNR, may intubate.” Plan for home hospice. Only receiving steroids, pain meds, and bowel regimen. Keeping foley for comfort despite risk of infection. |
|  |  | Transferred to inpatient hospice. "Suspected wall port may be seeded with bacteria as patient continued to experience rigors and expiratory wheezes when IV medications were administered through port. Given that patient had transitioned to comfort focused care, port removal was deferred as it's an invasive procedure." |
| Abbreviations: AKI, acute kidney injury; DNI, do not intubate; DNR, do not resuscitate; ED, emergency department; ICU, intensive care unit; IV, intravenous<br>*If coders were having difficulty distinguishing between “life extension” and “maintain or improve function” for a particular epoch, they were advised to use code status to distinguish between the two categories as follows: no limitations on resuscitation was considered more consistent with “life extension,” whereas documented plans to limit resuscitation (DNR and/or DNI) were considered more consistent with “maintain or improve function.” |  |  |

While any epoch could include individual therapies that fell into multiple categories, reviewers’ were instructed to assign a single best-fit overall category for that time period.

Specific considerations were given to time-limited trials of critical care or other invasive interventions such that if the intent was to identify and treat reversible issues to regain some baseline function then it would be considered consistent with “maintain or improve function” rather than “life-extension.” Similarly, code status was intentionally not considered a primary defining feature of any category. However, reviewers could use code status to help distinguish between the categories “life-extension” (e.g., full code) and “maintain or improve function” (e.g., do-not-attempt resuscitation (DNAR) or do-not-intubate (DNI)) when the intent of the care received was otherwise unclear.

Reviewers additionally reported their confidence in the care received assessment for each epoch using a 5-point Likert scale (1=not confident at all, 3=moderately confident, 5=very confident). They also identified prespecified diagnostic or therapeutic interventions within each epoch that assisted in their care received assessment (Table 2). Disagreements were first discussed by the two reviewers, and only if consensus could not be reached, physician investigators with expertise in critical care and palliative care (CA and KC) performed an independent review and adjudication.

**Table 2.** Specific diagnostic and therapeutic interventions assessed during chart review and frequency of occurrence.

| **Intervention** | **Frequency (%)**<br>**across 398**<br>**epochs** | **Frequency (%)**<br>**across 109**<br>**patients** |
| --- | --- | --- |
| Invasive diagnostic or therapeutic minor procedure* | 144 (36) | 80 (73) |
| Physical and/or occupational therapy (inpatient or outpatient) | 115 (29) | 74 (68) |
| DNR or DNI order | 65 (16) | 54 (50) |
| Admission to intensive care unit | 54 (14) | 43 (39) |
| Hospice enrollment | 51 (13) | 38 (35) |
| Chemotherapy | 48 (12) | 31 (28) |
| Invasive palliative procedure** | 43 (11) | 26 (24) |
| Intubation with mechanical ventilation | 38 (10) | 19 (17) |
| Comfort-care only order | 35 (9) | 31 (28) |
| Admission to short-term skilled nursing facility or acute rehabilitation center | 30 (8) | 24 (22) |
| Dialysis | 25 (6) | 13 (12) |
| Home palliative care consult | 24 (6) | 16 (15) |
| Surgical procedure | 22 (6) | 22 (20) |
| Palliative extubation/ventilator withdrawal | 6 (2) | 6 (6) |
| ACLS/CPR | 6 (2) | 6 (6) |
| Deactivation of ICD/pacemaker | 1 (0.3) | 1 (1) |
| * Invasive diagnostic or therapeutic minor procedures included: central venous catheter, feeding tube, thoracentesis, paracentesis, lumbar puncture, extracorporeal membrane oxygenation, chest tube insertion, biopsy, bronchoscopy, interventional radiology procedure<br>** Invasive palliative procedures included: thoracentesis, paracentesis, pleural catheter placement, nerve block, venting gastrostomy tube placement, lumbar puncture to relieve elevated intracranial pressure<br>ACLS, advanced cardiac life support; CPR, cardiopulmonary resuscitation; DNR, do not resuscitate; DNI, do not intubate; ICD, internal cardiac defibrillator |  |  |

#### 2.3.3 Goal-concordant Care Outcome

A goal-concordant epoch was defined as alignment between the GOC and care received assessments (e.g., both GOC and care received categorized as “life-extension”), and neither was classified as “unclear.” A goal-discordant epoch was defined as misalignment between the GOC and care received assessments (e.g., GOC categorized as “life-extension,” and care received categorized as “comfort-focused”), and neither was classified as “unclear”. Finally, uncertain concordance was defined when any GOC discussion was absent, or when GOC or care received were classified as “unclear.”

### 2.4 Statistical Analyses

We calculated the raw agreement percentage between reviewers for classification of care received and measured interrater reliability using Cohen’s kappa statistic. For assessments of GCC delivery, the primary unit of analysis was an epoch between GOC discussions. The association between each GOC category and receipt of goal-discordant care was assessed using logistic regression. Statistical significance was set at an alpha of 0.05 for 2-tailed tests. Statistical analyses were conducted using Stata v17 (College Station, TX).

## 3 Results

### 3.1 Patient Cohort and Documented GOC Discussions

Baseline demographic and clinical characteristics of the 109 unique patients in this cohort as well as the classification of GOC discussions have been reported previously.^20^ Briefly, 49 patients (45%) had a documented GOC discussion in the six months prior to the index hospital admission, thereby defining their baseline GOC. From the index hospital admission date through six months of follow-up, there were 289 GOC discussions among 83 (76%) patients. This resulted in a total of 398 epochs among the full cohort (Figure 2).

**Figure 2.**
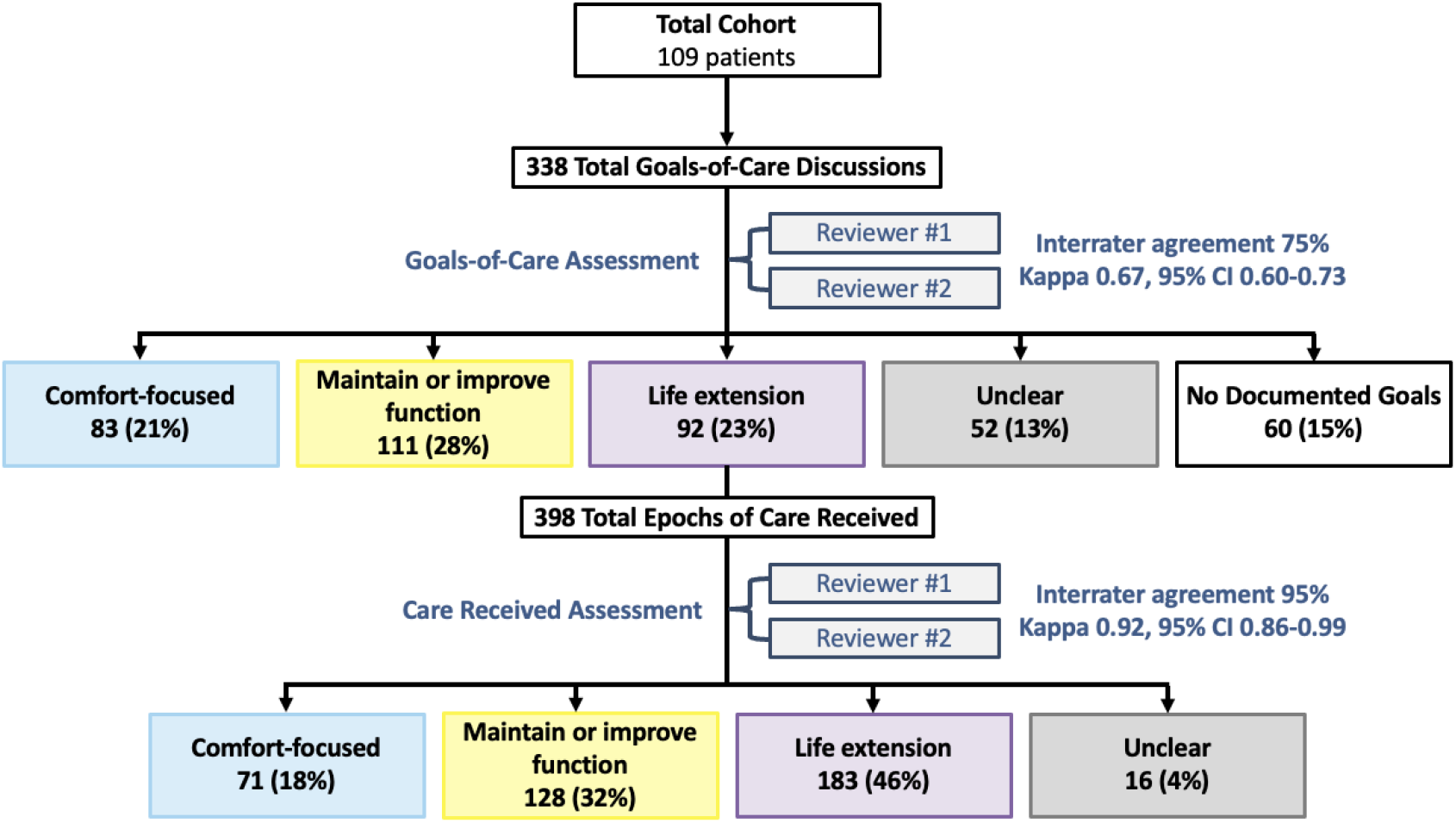
Study flow and classification of goals-of-care and care received.

### 3.2 Categorization of Care Received

Inter-rater reliability for classifying care received was almost perfect (95% interrater agreement; Cohen’s kappa=0.92; 95% CI, 0.86-0.99). Overall, the most common category of care received was “life-extension” (N=183, 46%), followed by “maintain or improve function” (N=128, 32%), “comfort-focused” (N=71, 18%), and “unclear” (N=16, 4%) (Figure 3). The median confidence score for assessing care received across all epochs was 4 (IQR 3, 5), with roughly equal proportions of epochs scored as moderately confident, considerably confident, and very confident (27%, 29%, and 30%, respectively). Reviewers were more often moderately to very confident when categorizing care received as “comfort-focused” compared to “life-extension” or “maintain or improve function” (93% vs 85% and 82%, respectively, p=0.01). Excerpts of clinical text representative of each care received category are provided in Table 1. Frequency of specific treatments or procedures are shown in Table 2.

**Figure 3.**
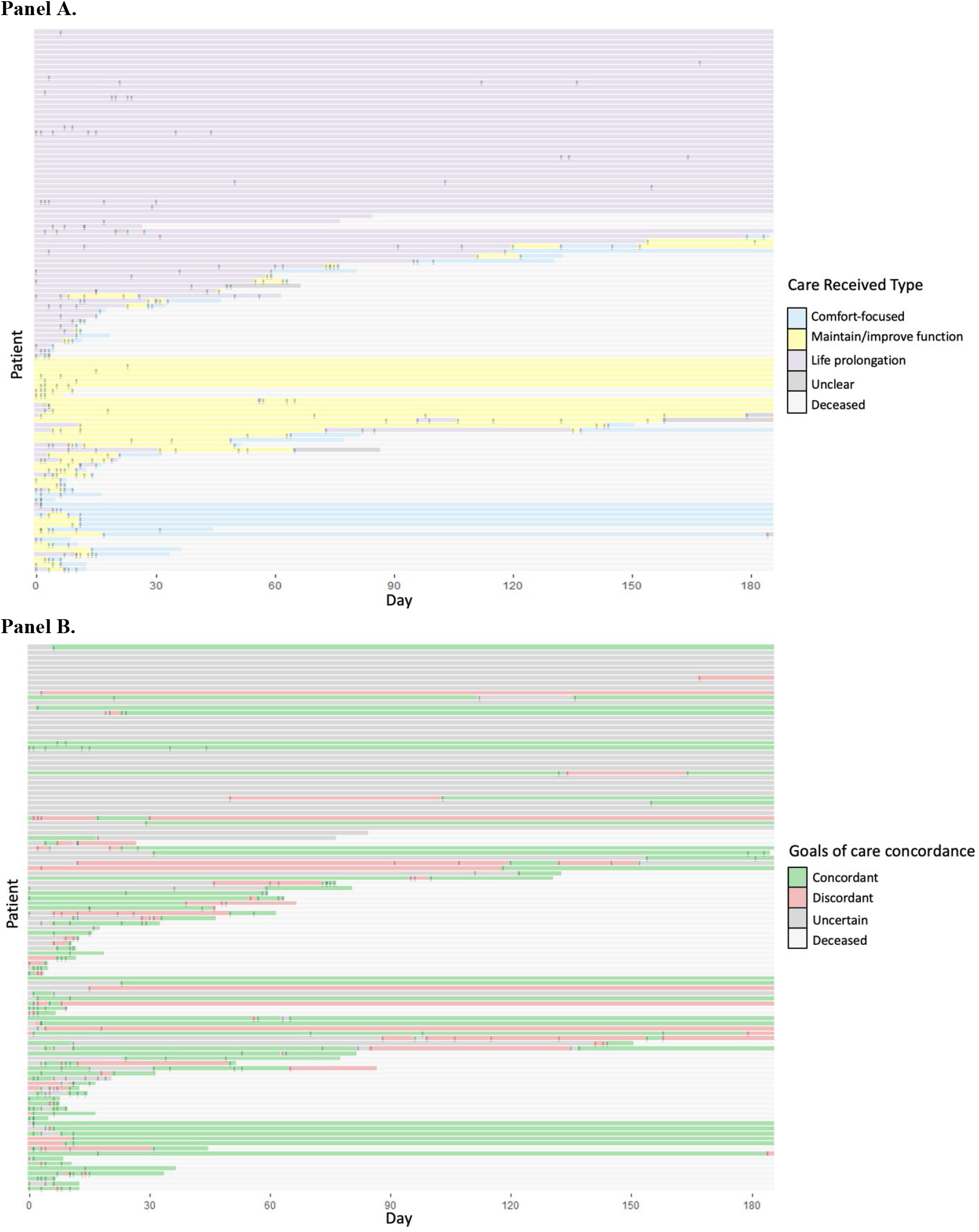
Tile plots showing frequency and classification of (**Panel A**) patients’ care received and (**Panel B**) goal-concordant, goal-discordant, or uncertain care over time. Made with R Core Team (2021). Ggplot. https://www.R-project.org/ Each patient is represented on the y-axis. Gray dashes denote individual goals-of-care discussions for each patient during the study period.

### 3.3 Goal-Concordant, Goal-Discordant, and Uncertain Care

GCC was identified in 198 (50%) of the total 398 epochs observed across 109 unique patients (Figure 3). Uncertain concordance was identified in 112 (28%) total epochs. Of these, the uncertainty was due to absence of a documented GOC discussion for 60 (54%) and GOC being classified as “unclear” in the remaining 52 epochs (46%). GCC was identified in 88 (22%) epochs. Receipt of goal-discordant care was most common when the documented goals were categorized as “maintain or improve function” as compared to “life-extension” or “comfort-focused” goals (40%, versus 24% and 27%, respectively, p=0.01).

### 3.4 Patient-Level, Epoch-Based Analyses

Patients had a median of 3 epochs (interquartile range (IQR) 2, 5) in the 6-month follow-up period. The median duration of each epoch was 7 days (IQR 2, 115). Patients received GCC in a median of 40% (IQR 0%, 75%) of epochs. Eighty (73%) patients received GCC during at least one epoch.

Among the 85 patients who had at least one documented GOC discussion, eight (9%) received GCC during every epoch; no patient characteristics were significantly associated with receiving 100% GCC. Among the fifty patients (46%) who died during the 6-month follow-up period, 42 (84%) were receiving GCC at the time of death, 5 (10%) care of uncertain concordance, and 3 (6%) goal-discordant care.

No patient received goal-discordant care for the entire six-month follow-up. Forty-eight (44%) patients received goal-discordant care during at least one epoch. The median proportion of epochs classified as goal-discordant care per patient was 0% (IQR 0%, 33%). Receipt of goal-discordant care was more common among patients admitted to a medical service compared to a surgical service (48% vs 16%, p=0.01) and more common among patients with a baseline diagnosis of metastatic cancer compared to those without (58% vs 32%, p<0.01).

Eighty (73%) patients received care of uncertain concordance during at least one epoch. The median proportion of epochs identified as uncertain concordance per patient was 33% (IQR 0%, 67%). Receipt of care of uncertain concordance was less common among patients who received a palliative care consultation during the index admission compared to those who did not (58% vs 83% vs p=0.004) and less common among patients with a baseline diagnosis of metastatic cancer compared to those without (60% vs 85%, p=0.004).

Among the 83 (76%) patients whose follow-up time spanned more than 1 epoch, 76 (92%) patients experienced epochs with variable care concordance (Figure 3). Of the 68 (62%) patients whose initial epochs were classified as uncertain concordance, 43 (63%) patients subsequently experienced an epoch in which concordance was either concordant (n=26, 60%) or discordant (n=17, 40%) (Figure 3). Of the 41 (38%) patients whose initial epochs were classified as either concordant or discordant, 12 (29%) patients experienced a subsequent epoch classified as uncertain concordance.

## 4 Discussion

In this retrospective chart review study, we identified clinically relevant epochs based on when a patient’s goals were documented by their clinicians, and then classified care received during that time as goal-concordant, goal-discordant, or of uncertain concordance. Our findings show promise for an entirely EHR-based method to reliably identify whether GCC was received or not among hospitalized patients at high risk of six-month mortality. This approach avoids the recall, social desirability, and selection biases that have plagued patient-, caregiver-, or clinician-reported assessments of goal-concordant or discordant care. Importantly, this approach relies on the practical data clinicians have available when caring for seriously ill patients and their families.

Our findings also demonstrate that GCC is not a static measure, thus highlighting the need to evaluate this outcome longitudinally. For example, nearly three-fourths of patients received GCC during at least one epoch and almost half also received goal-discordant care during at least one epoch. Assessing goal-concordance in longitudinal data has previously been limited by temporal gaps between goals and treatment,^6^ recognizing that goals can change over time.^28^ We overcame this limitation by capturing the multiple GOC discussions that tend to naturally occur in serious illness over time. And the median duration of an epoch among this cohort was one week, suggesting that most GCC assessments were based on the current GOC.

Another prior challenge to assessing GCC has been accurately categorizing care received.^6^ A narrow focus on specific treatments could lead to misclassification without considering the clinical context and intent in which they are provided. For this reason, we did not assign specific treatments to a category of care received, but rather we approached the assessment of care received holistically by using all of the available clinical note data. The same treatment for patients with different goals may be goal-concordant in both cases, as we found in this study (e.g., antibiotics to treat symptomatic cystitis may be consistent with “comfort-focused care” for one patient, while antibiotics to treat septic shock may be consistent with “life-extension” or “maintain or improve function” for another).

These findings also suggest that GOC and care received categories may not need to be perfectly aligned to indicate GCC. Indeed, nearly all care that is consistent with a goal to “maintain and improve function” may also be consistent with the goal of “life-extension,” because after all, one must be alive in order to function. This concept is supported by our prior work showing that patients and families often do hold multiple GOC concurrently.^20^ However, there is likely a point at which the specific treatment being considered primarily serves one goal at the expense of another (e.g., resuscitation in the event of cardiac arrest). More work is needed to understand how to handle overlapping GOC in the specification of GCC to account for the nuanced reality of serious illness care.

Uncertainty in concordance of care received was due to either lack of a documented GOC discussion or GOC truly being unclear to the physician reviewers. While some unclear GOC likely represent the real uncertainty that many patients and families experience in serious illness decision-making,^20^ using note templates with prompts and communication training could improve the clarity of GOC documentation. We found an association between receipt of goal-discordant care and being admitted to a medical compared to surgical service. We did not distinguish whether admissions were planned or unplanned, so it is possible that a greater proportion of surgical admissions were for planned procedures for which there was greater opportunity to consider and ensure that treatment would align with patient goals. Goal-discordant care was also noted to be more common when goals were “maintain or improve function” compared to “life-extension” or “comfort-focused.” These findings may be explained by the clinical moment that often prioritizes aggressive, life-prolonging treatments unless or until patients or families clearly state a desire for comfort-focused care.^29^ While there are active efforts to encourage clinicians to consider when comfort-focused care might be appropriate,^7^ and EHR order-sets even exist to support such care,^30^ there has been less attention on identifying and supporting goals focused on function. Confirmation of these findings in a larger, more diverse sample is needed.

There are several notable strengths of this study. First, we broadly defined serious illness to promote generalizability of our approach to measuring GCC among a typical hospitalized population. Second, our novel approach to use naturally occurring GOC discussions over time to define clinical epochs revealed the dynamic nature of GCC delivery and the importance of a longitudinal assessment. Third, we employed a multitude of chart review methods that mitigated reviewer bias to promote veracity of our findings. Finally, demonstrating feasibility of this novel EHR-based approach to identify GCC serves as important proof-of-concept for the promise of natural language processing and machine learning methods to augment widespread applicability.^18,21,22^

The findings from the study must also be interpreted considering several limitations. First, we conducted this work in a single health system. Local culture can influence clinicians’ GOC communication and documentation as well as serious illness treatment practices, thus validation of this approach in another health system is necessary. Second, we relied on reviewers’ interpretation of the primary intent of the care patients’ received from EHR documentation. A prospective validation study would allow for a comparison of the care received classification with the actual intent of care from the treatment team in real time. Third, this study was conducted without patient or family input. An important step before optimizing the pragmatism of this novel framework will be to validate the measurement approach with key stakeholders, including patients, families, and clinicians from diverse settings. Finally, manual chart review by clinician reviewers was necessary for this feasibility study, however, it is highly resource-intensive and future efforts in larger cohorts will need to leverage artificial intelligence methods for practicability.

## 5 Conclusions

Measurement of GCC using clinician review of EHR data is feasible on a small scale, and revealed specific opportunities to improve serious illness communication and documentation to avoid goal-discordant care. Incorporating patient- and family perspectives could enhance the patient-centeredness of an EHR-based GCC outcome measure that could then be externally validated, optimized, and automated for widespread clinical and research use.

## Data Availability

All data produced in the present study are available upon reasonable request to the authors.

## Notes

**Funding:** Dr. Auriemma is supported by a National Institute of Health, National Heart, Lung, and Blood Institute (NHLBI) Grant (K23-HL163402) and a National Institute of Health Loan Repayment Program Award (L30HL154185). Dr. Song is supported by NHLBI Grant T32HL098054. Dr. Weissman is supported by R03HL171424 and R35GM155262. Dr. Halpern is supported by NHLBI Grant 2K24 HL143289. Dr. Courtright is supported by National Institute on Aging (NIA) Grant R01 AG073384. The authors had full independence in designing the study, interpreting the data, writing, and publishing the report. The content is solely the responsibility of the authors and does not necessarily represent the official views of the University of Pennsylvania or National Institutes of Health.

### Competing Interest Statement

The authors have declared no competing interest.

### Funding Statement

Dr. Auriemma is supported by a National Institute of Health, National Heart, Lung, and Blood Institute (NHLBI) Grant (K23-HL163402) and a National Institute of Health Loan Repayment Program Award (L30HL154185). Dr. Song is supported by NHLBI Grant T32HL098054. Dr. Weissman is supported by R03HL171424 and R35GM155262. Dr. Halpern is supported by NHLBI Grant 2K24 HL143289. Dr. Courtright is supported by National Institute on Aging (NIA) Grant R01 AG073384. The authors had full independence in designing the study, interpreting the data, writing, and publishing the report. The content is solely the responsibility of the authors and does not necessarily represent the official views of the University of Pennsylvania or National Institutes of Health.

### Author Declarations

The Institutional Review Board of the University of Pennsylvania approved this study with a waiver of informed consent under protocol number 849694.

## References

1. Bernacki RE, Block SD. Communication about serious illness care goals: a review and synthesis of best practices. JAMA Intern Med. 2014;174(12):1994–2003. doi:10.1001/JAMAINTERNMED.2014.5271

2. Dy SM, Kiley KB, Ast K, et al. Measuring what matters: top-ranked quality indicators for hospice and palliative care from the American Academy of Hospice and Palliative Medicine and Hospice and Palliative Nurses Association. J Pain Symptom Manage. 2015;49(4):773–781. doi:10.1016/J.JPAINSYMMAN.2015.01.012

3. Turnbull AE, Hartog CS. Goal-concordant care in the ICU: a conceptual framework for future research. Intensive Care Med. 2017;43(12):1847–1849. doi:10.1007/s00134-017-4873-2

4. Sanders JJ, Curtis JR, Tulsky JA. Achieving Goal-Concordant Care: A Conceptual Model and Approach to Measuring Serious Illness Communication and Its Impact. J Palliat Med. 2018;21(S2):S17-S27. doi:10.1089/jpm.2017.0459

5. Halpern SD. Goal-concordant care — Searching for the Holy Grail. New England Journal of Medicine. 2019;381(17):1603–1606. doi:10.1056/NEJMp1908153

6. Ernecoff NC, Mph KLW, Bennett A V, Hanson LC, Bennett A V, Hanson LC. Measuring Goal-Concordant Care in Palliative Care Research. J Pain Symptom Manage. 2021;62(3):e305–e314. doi:10.1016/j.jpainsymman.2021.02.030

7. Zimmerman JJ, Harmon LA, Smithburger PL, et al. Choosing Wisely For Critical Care: The Next Five. Crit Care Med. 2021;49(3):472–481. doi:10.1097/CCM.0000000000004876

8. American Society for Radiation Oncology. Five things physicians and patients should question. Choosing Wisely: An Initiative of the ABIM Foundation. Published online 2013. Accessed April 9, 2024. https://www.oregonclinic.com/wp-content/uploads/2023/07/ASTRO-Choosing-Wisely-five-things-to-question.pdf

9. Commission on Cancer. Five things physicians and patients should question. Choosing Wisely: An Initiative of the ABIM Foundation. Published online 2012. Accessed April 9, 2024. https://www.facs.org/media/wegncq3f/coclist.pdf

10. Dying in America: Improving Quality and Honoring Individual Preferences Near the End of Life. Dying in America: Improving Quality and Honoring Individual Preferences Near the End of Life. Published online April 19, 2015:1–638. doi:10.17226/18748

11. Sudore RL, Heyland DK, Lum HD, et al. Outcomes That Define Successful Advance Care Planning: A Delphi Panel Consensus. J Pain Symptom Manage. 2018;55(2):245-255.e8. doi:10.1016/j.jpainsymman.2017.08.025

12. Sanders JJ, Curtis JR, Tulsky JA. Achieving Goal-Concordant Care: A Conceptual Model and Approach to Measuring Serious Illness Communication and Its Impact. J Palliat Med. 2018;21(S2):S17-S27. doi:10.1089/jpm.2017.0459

13. Parks Taylor S, Kowalkowski MA, Courtright KR, et al. Deficits in Identification of Goals and Goal-Concordant Care After Sepsis Hospitalization. An Official Publication of the Society of Hospital Medicine Journal of Hospital Medicine ®. 2021;16(11). doi:10.12788/jhm.3714

14. Landrum KKM, Ross T, Andiel C, et al. Meaningful measures for advanced cancer: Building a measure set to promote goal-concordant care. 101200/JCO20224028_suppl210. 2022;40(28_suppl):210–210. doi:10.1200/JCO.2022.40.28_SUPPL.210

15. Modes ME, Heckbert SR, Engelberg RA, Nielsen EL, Curtis JR, Kross EK. Patient-reported receipt of goal-concordant care among seriously ill outpatients - prevalence and associated factors. J Pain Symptom Manage. 2020;60(4):765. doi:10.1016/J.JPAINSYMMAN.2020.04.026

16. Modes ME, Engelberg RA, Nielsen EL, et al. Seriously Ill Patients’ Prioritized Goals and Their Clinicians’ Perceptions of Those Goals. J Pain Symptom Manage. 2022;64(4). doi:10.1016/J.JPAINSYMMAN.2022.06.004

17. Weissenbacher D, Courtright K, Rawal S, et al. Detecting goals of care conversations in clinical notes with active learning. J Biomed Inform. 2024;151. doi:10.1016/J.JBI.2024.104618

18. Lee RY, Brumback LC, Lober WB, et al. Identifying Goals of Care Conversations in the Electronic Health Record Using Natural Language Processing and Machine Learning. J Pain Symptom Manage. 2021;61(1):136-142.e2. doi:10.1016/J.JPAINSYMMAN.2020.08.024

19. Randall Curtis JR, Sathitratanacheewin S, Starks H, et al. Using Electronic Health Records for Quality Measurement and Accountability in Care of the Seriously Ill: Opportunities and Challenges. J Palliat Med. 2018;21(S2):S52–S60. doi:10.1089/JPM.2017.0542

20. Auriemma CL, Song A, Walsh L, et al. Classification of Documented Goals of Care Among Hospitalized Patients with High Mortality Risk: a Mixed-Methods Feasibility Study. J Gen Intern Med. Published online 2024. doi:10.1007/S11606-024-08773-Z

21. Chan A, Chien I, Moseley E, et al. Deep learning algorithms to identify documentation of serious illness conversations during intensive care unit admissions. 101177/0269216318810421. 2018;33(2):187–196. doi:10.1177/0269216318810421

22. Avati A, Jung K, Harman S, Downing L, Ng A, Shah NH. Improving palliative care with deep learning. BMC Med Inform Decis Mak. 2018;18(4):55–64. doi:10.1186/S12911-018-0677-8/TABLES/5

23. Lee RY, Kross EK, Torrence J, et al. Assessment of Natural Language Processing of Electronic Health Records to Measure Goals-of-Care Discussions as a Clinical Trial Outcome. JAMA Netw Open. 2023;6(3):E231204. doi:10.1001/JAMANETWORKOPEN.2023.1204

24. Nastasi AJ, Courtright KR, Halpern SD, Weissman GE. A vignette-based evaluation of ChatGPT’s ability to provide appropriate and equitable medical advice across care contexts. Sci Rep. 2023;13(1). doi:10.1038/S41598-023-45223-Y

25. Martin JA, Crane-Droesch A, Lapite FC, et al. Development and validation of a prediction model for actionable aspects of frailty in the text of clinicians’ encounter notes. J Am Med Inform Assoc. 2021;29(1):109–119. doi:10.1093/JAMIA/OCAB248

26. Courtright KR, Chivers C, Becker M, et al. Electronic Health Record Mortality Prediction Model for Targeted Palliative Care Among Hospitalized Medical Patients: a Pilot Quasi-experimental Study. J Gen Intern Med. 2019;34(9):1841–1847. doi:10.1007/s11606-019-05169-2

27. Secunda K, Wirpsa MJ, Neely KJ, et al. Use and Meaning of “Goals of Care” in the Healthcare Literature: a Systematic Review and Qualitative Discourse Analysis. J Gen Intern Med. 2020;35(5):1559–1566. doi:10.1007/S11606-019-05446-0

28. Auriemma CL, Nguyen CA, Bronheim R, et al. Stability of end-of-life preferences: A systematic review of the evidence. JAMA Intern Med. 2014;174(7):1085–1092. doi:10.1001/jamainternmed.2014.1183

29. Kruser JM, Cox CE, Schwarze ML. Clinical Momentum in the Intensive Care Unit. A Latent Contributor to Unwanted Care. Ann Am Thorac Soc. 2017;14(3):426–431. doi:10.1513/ANNALSATS.201611-931OI

30. Bender MA, Hurd C, Solvang N, Colagrossi K, Matsuwaka D, Curtis JR. A New Generation of Comfort Care Order Sets: Aligning Protocols with Current Principles. J Palliat Med. 2017;20(9):922–929. doi:10.1089/JPM.2016.0549

